# AgeNet-SHAP: An explainable AI approach for optimally mapping multivariate regional brain age and clinical severity patterns in Alzheimer’s disease

**DOI:** 10.1101/2025.02.28.25323097

**Authors:** Gauri Darekar, Taslim Murad, Hui-Yuan Miao, Deepa S. Thakuri, Alzheimer’s Disease Neuroimaging Initiative, Ganesh B. Chand

**Affiliations:** Department of Radiology, Mallinckrodt Institute of Radiology, Washington University School of Medicine, St. Louis, MO, USA; Institute for Informatics, Data Science and Biostatistics, Washington University School of Medicine, St. Louis, MO, USA; University of Missouri, School of Medicine, Columbia, MO, USA; Imaging Core, Knight Alzheimer Disease Research Center, Washington University School of Medicine, St. Louis, MO, USA; Institute of Clinical and Translational Sciences, Washington University School of Medicine, St. Louis, MO, USA; NeuroGenomics and Informatics Center, Washington University School of Medicine, St. Louis, MO, USA

**Keywords:** Alzheimer’s Disease (AD), Mild Cognitive Impairment (MCI), Brain Age, Deep Learning (DL), Machine Learning (ML), Feature Importance, SHapley Additive exPlanations (SHAP)

## Abstract

Age is a significant risk factor for mild cognitive impairment (MCI) and Alzheimer’s disease (AD) and identifying brain age patterns is critical for comprehending the normal aging and MCI/AD processes. Prior studies have widely established the univariate relationships between brain regions and age, while multivariate associations remain largely unexplored. Herein, various artificial intelligence (AI) models were employed to perform brain age prediction using an MRI dataset (n=668). Then the optimal AI model was integrated with the Shapley additive explanations (SHAP) feature importance technique to identify the significant multivariate brain regions involved in this prediction. Our results indicated that the deep learning model (referred to as AgeNet) tremendously outperformed the conventional machine learning models for brain age prediction, and AgeNet integrated with SHAP (referred to as AgeNet-SHAP) identified all ground-truth perturbed regions as key predictors of brain age in semi-simulation, proved the validity of our methodology. In the experimental dataset, compared to cognitively normal (CN) participants, MCI exhibited moderate differences in brain regions, whereas AD had highly robust and widely distributed regional differences. The individualized AgeNet-SHAP regional features further showed associations with clinical severity scores in the AD continuum. These results collectively facilitate data-driven predictive modelling approaches for disease progression, diagnostics, prognostics, and personalized medicine efforts.

## 1. Introduction

Alzheimer’s disease (AD) is the most prevalent neurodegenerative disorder, characterized by neural degeneration and structural changes in the brain. It is a leading cause of dementia, representing a significant global health challenge, with over 55 million individuals affected worldwide [1]. In the United States alone, over 6.9 million adults aged 65 and older are affected, and projections indicate that this number will double by 2050 [2]. This highlights the pressing need to develop effective preventive strategies and therapeutic interventions for AD, which requires a clear understanding of AD mechanisms. To fully uncover AD mechanisms, it is crucial to have a deeper understanding of the various facets of AD, including factors such as age and brain alterations.

Human brain changes with chronological aging process, and neurodegenerative diseases such as AD can disrupt the normal aging trajectory and accelerate such process [3, 4]. Brain age, a reliable biomarker for neurodegenerative diseases derived from neuroimaging data, quantifies this accelerated aging process. The difference between chronological age and brain age is often referred to as the brain age gap, with a larger gap being associated with more significant cognitive impairment [5]. Thus, understanding brain alterations accompanying brain aging in healthy and MCI/AD populations could provide valuable insights in the MCI/AD mechanisms.

Brain structural alterations can be assessed using neuroimaging markers [4, 6, 7], like magnetic resonance imaging (MRI) [4], which, for instance has demonstrated an association between gray matter volume reduction and increased age [8, 9]. The integration of neuroimaging with machine learning (ML) and artificial intelligence (AI) techniques has led to significant advancements in AD research [10-13]. However, the existing reports related to brain age in AD predominantly focus on characterizing univariate relationships between age and individual brain regions or tissue types. In reality, multiple brain regions show associations with age [14, 15] and work collaboratively in brain processes [7, 16, 17], hence highlighting the need to capture multivariate interactions between brain regions for precise understanding of brain aging mechanisms. Moreover, it is unknown whether multivariate regional patterns of brain age are associated with clinical severity in MCI/AD. Such multivariate predictive modelling can collectively aid in designing more effective targeted preventive and therapeutic strategies for AD.

In this study, we investigated the performance of several AI models, including conventional ML models and a deep learning (DL) model named AgeNet, for brain age prediction and integrated the optimal predictive model, which is the AgeNet model, with the Shapley additive explanations (SHAP) [18] feature importance technique, referred to as AgeNet-SHAP, to identify the hierarchy of multivariate associations between brain regions and brain age in health control and AD populations. We hypothesize that our approach can effectively capture the important regional hubs related to aging in health and AD populations, and a comparison between those two populations could shed light on the mechanisms of AD. In addition, we hypothesize that our approach could also illustrate the clinical severity relevance of those regions under the framework of AgeNet-SHAP.

## 2. Methods and Materials

### 2.1. Datasets

T1-weighted MR images of 668 participants (55.1-91.5 years old, with a mean of 72.83 and standard deviation of 7.34 years old; 308 females) at their baseline visit from the Alzheimer’s Disease Neuroimaging Initiative (ADNI)-2 database [19] were used for this study. There are 187 cognitively normal (CN) individuals, and 481 MCI/AD patients based on record of their baseline visit. Using the Multi-atlas region segmentation utilizing ensembles of registration techniques and parameter modification (MUSE) method [20], each MRI was segmented into 145 distinct regions of interest (ROIs), encompassing gray matter, white matter, and cerebrospinal fluid areas. The volumetric measurement of each ROI was extracted and then corrected for the site and sex influence by employing an existing regression-based harmonization technique [17, 21]. Then, the ROI volumes for each participant are normalized by dividing each ROI by the sum of all ROIs. Normalization is crucial for removing head size differences and making the data more suitable for AI modeling. We utilized the normalized harmonized ROIs for our analysis.

#### 2.1.1 Simulated Data

We formulated semi-simulated data to validate the ability of AgeNet-SHAP approach for capturing the hierarchical multivariate relationships between brain regions and brain age. The aim of the simulation analysis is to establish arbitrary multivariate correlations between brain volumetric measures and brain age, and to evaluate whether our methodology can accurately detect these complex relationships. The semi-simulated data was created using only CN participants (n=187), which can enable the preservation of normal brain variations. To generate the data, we chose the top 10 ROIs having strongest negative correlation with age. Given that brain volumetric measurements decrease with age, negative correlations are considered here. The ROIs selected for perturbation are the right thalamus proper, left superior occipital gyrus, left planum temporale, right planum polare, right planum temporale, right posterior insula, left accumbens area, left anterior limb of internal capsule, left planum polare, and right anterior limb of internal capsule. For 40% participants we reduced the volumes of the selected ROIs by a factor between 30-50% and increased age by a factor between 10-30%. These factors were chosen from a random uniform distribution. The perturbations retain the maximum negative correlation between age and the selected ROIs, while maintaining suboptimal correlations for other ROIs.

#### 2.1.2 Experimental Data

We utilized the 668 MRI data from ADNI cohort as the experimental data. Among MCI/AD participants, 173 were labelled as early MCI (EMCI), 152 as late MCI (LMCI), 6 as MCI, and 150 as AD. The data also included the chronological age of each participant recorded on the MRI date. We utilized the normalized harmonized brain volumetric changes expressed through ROIs to predict the age, and this predicted age is referred to as brain age. Additionally, the Clinical Dementia Rating Scale Sum of Boxes (CDR-SB) [22, 23] scores of all participants are also collected to investigate the clinical severity processes. For each subject, the CDR-SB closest to the MRI date was selected.

### 2.2. Brain age prediction via AI models

We employed three ML models and one DL model to predict brain age. Then the optimal performing model was combined with SHAP to identify the significant multivariate brain regional hubs towards the brain age prediction.

The ML models used in this study include Lasso regression (LR) [24, 25], Ridge regression (RR) [26, 27], and support vector regression (SVR) [28]. These models were selected for their effectiveness in handling high-dimensional data, with SVR being particularly useful due to its non-linear capabilities. The hyperparameters for each model were optimized using the grid search approach based on the negative mean absolute error scoring metric. For LR and RR, the best alpha values were chosen within the ranges of 0.15-0.35 with an increment of 0.05 and 150-250 with an increment of 10, respectively. For SVR, the Radial Basis Function (RBF) kernel was employed, and the gamma, epsilon, and C hyperparameters were tuned across the following values: gamma for n=−12 to −2 with an increment of 1, epsilon for n=−7 to 1 with an increment of 1, and C for n=−1 to 4 with an increment of 1. These models were trained following a 10-fold stratified cross-validation (CV) procedure, where the data was spitted into training and testing set. The reported results were obtained from the test data. Note that, the test data is distinct from train data and is not used in training the models.

Given the superior ability of DL models over conventional ML models to capture complex non-linear relationships, we tested brain age prediction using a basic deep neural network (DNN). Considering the limitations of our dataset (i.e., small sample size), this model is the most suitable, as it provides promising and generalized results while minimizing overfitting due to its architectural simplicity. The DL model, named AgeNet, consists of five layers for estimating brain age. It has four hidden dense layers followed by a final output dense layer. The hidden layers contain decreasing units in the sequence of 256, 128, 64, and 32, each utilizing a rectified linear unit (ReLU) activation function, followed by a batch normalization and a dropout layer with a dropout rate of 0.4. The final output layer features a single unit with a linear activation function to predict the brain age value. The training parameters contain mean absolute error (MAE) loss function, ADAM optimizer, 0.01 learning rate, 64 batch size, and 300 epochs. The hyperparameters were tuned to optimize the model’s performance. This model is also trained using 10-fold stratified CV and the reported results are computed on the test data. The test data is not overlapped with the training data, hence ensuring the model’s generalizability.

Since the AgeNet model tends to perform better than the ML models for brain age prediction, we integrated it with SHAP method to identify the significant brain regional hubs involve in the brain age prediction. SHAP is a widely used technique for feature importance that accounts for interactions among features, unlike the permutation feature importance method [29]. It is also generally more stable and consistent than the Local Interpretable Model-agnostic Explanations (LIME) method [30]. The significance of a brain region (ROI) is determined based on its SHAP value, and this value is computed by considering the impact of the region on the final predictive output by adding and removing it from the input. The SHAP value for a region is obtained by averaging the absolute SHAP values of that feature for all participants. A higher average absolute SHAP indicates a higher significance of that feature for making the prediction. The equation [10] used to calculate the SHAP value for an input feature *r* is,

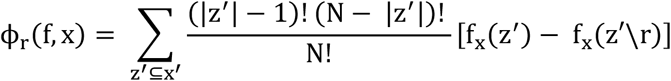

where *x* is the input ROIs, *y* is the output of the *f* AgeNet model. Let *x* be the simplified input which maps to the original input *x* using the function *x=h*_*x*_*(x), N* is a binary coalition vector, and *z* is a binary coalition vector that represents a perturbed version of the input features. The overall workflow AgeNet-SHAP is illustrated in **Figure 1**. We computed the SHAP values on the test data for each fold during the 10-fold stratified CV.

**Figure 1:**
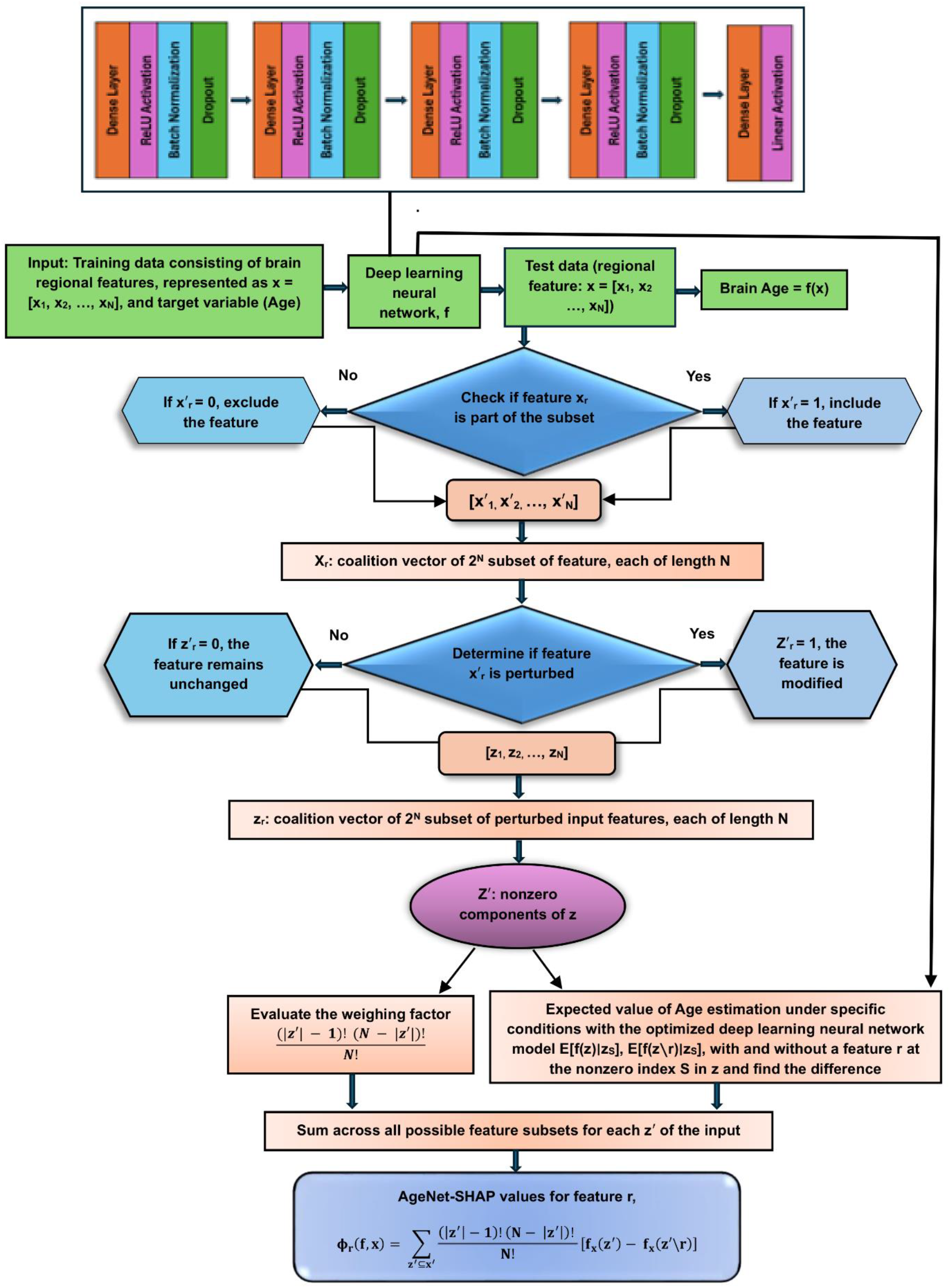
The schematic workflow diagram of the AgeNet-SHAP method.

For the robustness purposes, we ran our models’ multiple times (five times in this case) and averaged the results, where each run consists of 10-fold stratified CV training-testing process. Moreover, a model’s performance is evaluated by computing the Spearman correlation between the actual and predicted brain age within the test set, along with determining the significance of this correlation via the computed *p*-value, following the Spearman rank-order correlation coefficient method [31]. This is a non-parametric approach which does not make assumptions about the underlying data distribution. Then the *p*-values are mapped to false discovery rate (FDR) corrected *p*-values, as FDR-corrected *p*-values are best for validating the performance of various models because they help to control the rate of false positives when performing multiple comparisons, ensuring that observed significant correlations are more likely to reflect true effects rather than random variation. Additionally, to display the brain figures, they are overlayed on the standard MRI template in MNI space using the axial orientation. Additionally, we assessed brain structural differences between the CN group and individuals with MCI and AD using Cohen’s d [32], a widely recognized measure of effect size. Individualized AgeNet-SHAP regional values were used to compute Cohen’s d. Only ROIs with FDR-corrected *p*-values < 0.05 were considered statistically significant. Comparisons were made between the CN group and both MCI and AD groups to examine how brain alterations, in terms of multivariate regional associations with brain age, evolve with group-wise disease progression.

### 2.3. Clinical severity of AD

The clinical severity of MCI/AD was explored by investigating the correlations between the individualized AgeNet-SHAP regional features and the CDR-SB scores. The CDR-SB [22, 23] score ranges from 0 to 18, derived from the assessment of six cognitive domains (memory, orientation, judgment and problem-solving, community affairs, home and hobbies, and personal care), where higher scores indicate greater clinical severity. This analysis highlights the relevance of AgeNet-SHAP based generated features to determine the severity of MCI/AD.

## 3. Results

### 3.1. Simulated data results

The AgeNet model outperformed all the three conventional ML models for brain age prediction using the semi-simulated data, as given in **Table 1**. It achieved a correlation of 0.91 between the actual and predicted brain age, along with an FDR corrected *p*-value of 2.84E-73 (**Figure 2 (a)**), while the LR, RR, and SVR models obtained 0.769, 0.777, and 0.761 correlations, respectively. Note that, although the test loss score of AgeNet is higher than that of the ML models, it still yields an optimal correlation, indicating that the model captures non-linear complex relationships in the data more effectively than the ML models, even if it has not fully converged. Moreover, the AgeNet-SHAP was able to identify all perturbed brain regions to the significant brain regions for brain age prediction based on their SHAP values, as illustrated in **Figure 2 (b)**.

**Table 1.** The performance of Lasso regression (LR), Ridge regression (RR), support vector regression (SVR), and AgeNet for brain age predictions using the semi-simulated data.

| Models | | Test Loss | Spearman's Correlation | FDR-corrected $p$-value |
| --- | --- | --- | --- | --- |
| ML | LR | 5.26 | 0.769 | 7.63E-38 |
|  | RR | 5.18 | 0.777 | 5.09E-39 |
|  | SVR | 6.43 | 0.761 | 1.38E-36 |
| DL | AgeNet | 9.13 | 0.91 | 2.84E-73 |

**Figure 2:**
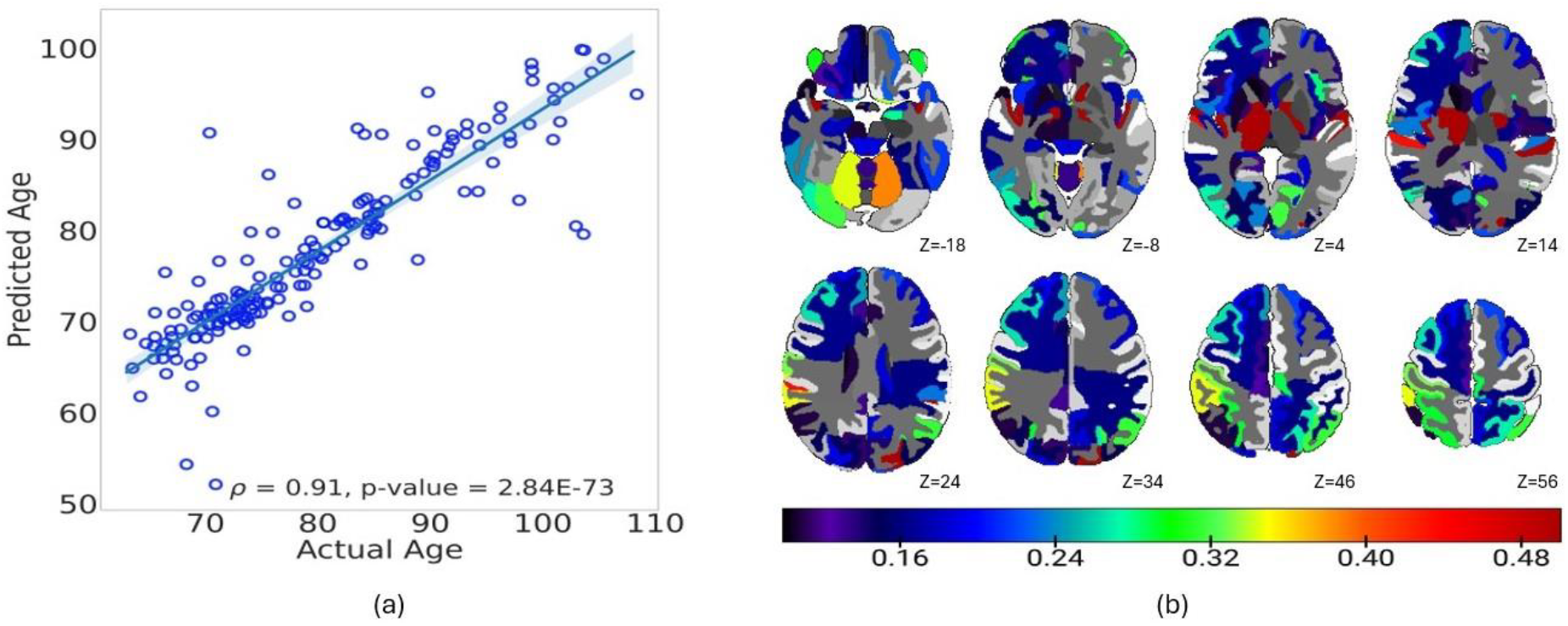
AgeNet-SHAP based brain age prediction using the semi-simulated data. **(a)** Spearman’s correlation between the actual and predicted brain age. **(b)** Significant brain regions identified by AgeNet-SHAP. The color bar indicates SHAP values. The results are averaged over all participants. z shows the brain slice position.

### 3.2. Experimental data results

The AgeNet model exhibited higher performance as compared to the conventional ML models for brain age prediction using the experimental dataset, as shown in **Table 2**. It obtained a correlation of 0.88 between the actual and predicted age (*p* = 1.14E-212, FDR corrected, **Figure 3 (a)**), along with yielding optimal test loss score. In contrast, the correlations achieved by LR, RR, and SVR are 0.655, 0.658, and 0.657, respectively. Additionally, the outperforming AgeNet model integrated with SHAP, AgeNet-SHAP, identified the significant brain regional hubs involved in brain age estimation across the CN, MCI, and AD groups, as illustrated in **Figure 3 (b), (c)**, and **(d)** respectively. In CN participants, the identified top significant brain regions are the left anterior limb of internal capsule, right anterior limb of internal capsule, left planum polare, left basal forebrain, and right posterior limb of internal capsule inc. cerebral peduncle. In the MCI category, the top key brain regions identified are the left anterior limb of internal capsule, right anterior limb of internal capsule, left planum polare, right posterior limb of internal capsule inc. cerebral peduncle, and third ventricle. For the individuals with AD, the top dominant brain regions involved in brain age prediction are the left anterior limb of internal capsule, right anterior limb of internal capsule, left planum polare, third ventricle, and brain stem. Moreover, **Figure 3 (e)** illustrates the Cohen’s d differences between the CN and MCI groups, and the regions with notable effect sizes include the right amygdala, left inferior temporal gyrus, left inferior lateral ventricle, left middle temporal gyrus, and left hippocampus. Similarly, **Figure 3 (f)** demonstrates the Cohen’s d differences between the CN and AD groups, and the regions with key effect sizes are the right amygdala, right inferior lateral ventricle, left middle temporal gyrus, left hippocampus, and left inferior lateral ventricle. Comparing **Figure 3 (e)** and **Figure 3 (f)**, AD patients show much bigger brain alterations than MCI patients when compared to CN group, both in the aspects of range and magnitude. For example, the right amygdala, left middle temporal gyrus, and left hippocampus regions in MCI portrayed moderate differences when compared to CN, while in AD they showed a much higher differences than CN.

**Table 2.** The performance of Lasso regression (LR), Ridge regression (RR), support vector regression (SVR), and AgeNet for brain age predictions using the experimental data.

| Models |  | Test Loss | Spearman's Correlation | *p*-value |
| --- | --- | --- | --- | --- |
| ML | LR | 4.42 | 0.655 | 5.87E-87 |
|  | RR | 4.40 | 0.658 | 5.20E-84 |
|  | SVR | 4.35 | 0.657 | 8.70E-84 |
| DL | AgeNet | 3.467 | 0.88 | 1.4E-212 |

**Figure. 3:**
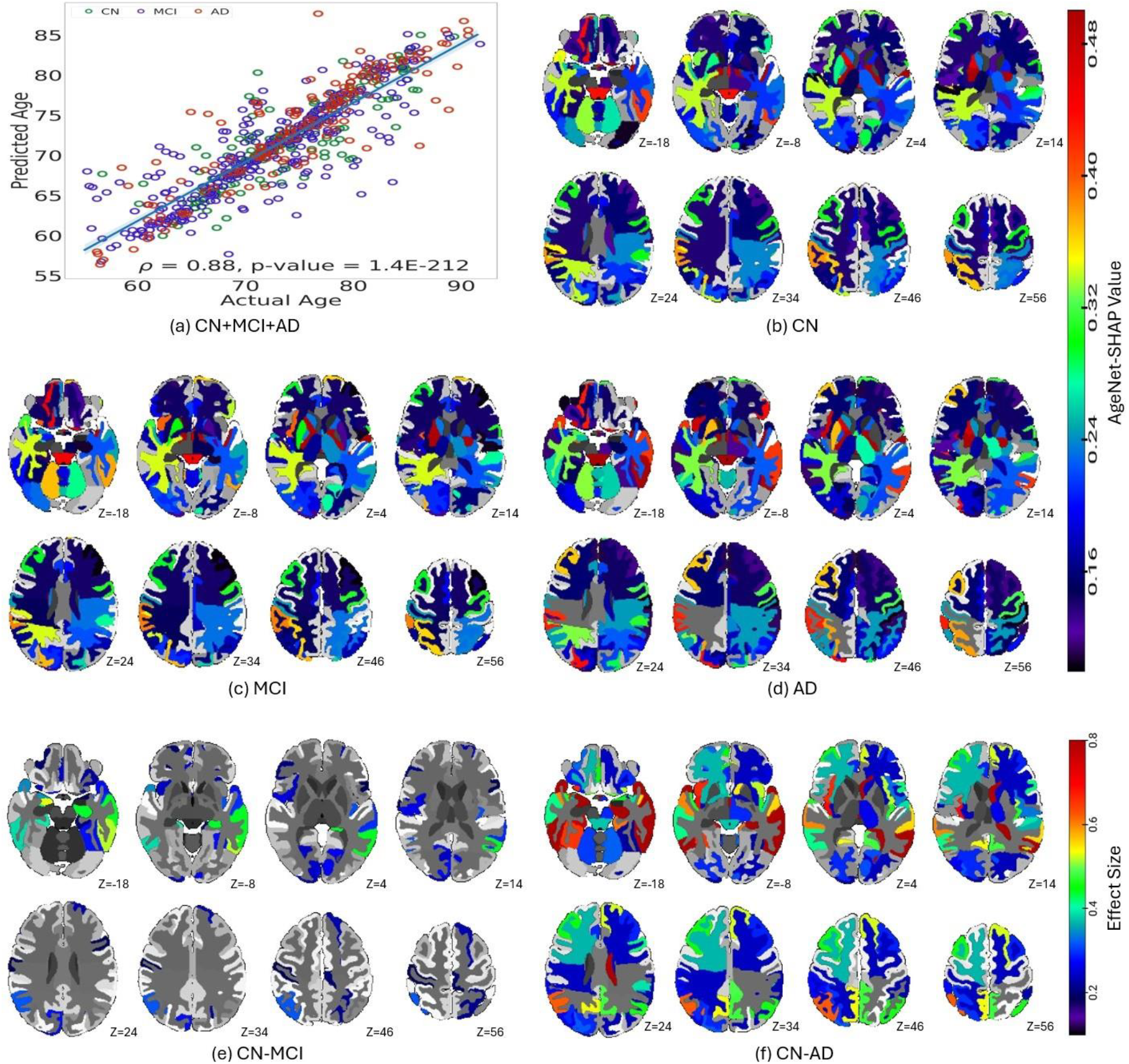
AgeNet-SHAP on experimental data. **(a)** Spearman’s correlation between the actual and predicted brain age for entire data. The individualized AgeNet-SHAP regional features were averaged within each group and significant brain regions are shown for **(b)** CN, **(c)** MCI, and (**d)** AD groups. Cohen’s d effect size group differences are presented for **(e)** CN – MCI and **(f)** CN – AD. The results are averaged over all participants. z shows the brain slice position. The gray areas show the background, and they are not correlated.

### 3.3. Clinical severity results

The multivariate regional AgeNet-SHAP features were studied in associations with the clinical severity scores in the AD continuum. The brain regions significantly associated with CDR-SB scores, based on individualized AgeNet-SHAP features, are depicted in **Figure 4** for the **(a)** combined (CN, MCI, and AD together), **(b)** CN, **(c)** MCI, and **(d)** AD groups. Only correlations with FDR-corrected *p*-values < 0.05 were considered significant and presented. Notable brain regions exhibiting positive correlations with CDR-SB in the combined group include the left precuneus, left superior frontal gyrus, left middle temporal gyrus, left inferior lateral ventricle, and right fornix. Conversely, prominent regions showing significant negative correlations with CDR-SB in the combined group include the left superior parietal lobule, left occipital fusiform gyrus, right basal forebrain, right parietal white matter, and left lateral orbital gyrus. Moreover, the regions with significant positive correlations in CN are the left parahippocampal gyrus and left lingual gyrus, while the regions with significant negative correlations are the left fornix, right middle temporal gyrus, left superior parietal lobule, and right basal forebrain. In MCI group, the dominant positive correlations are associated with the right precentral gyrus medial segment, right lateral orbital gyrus, right middle frontal gyrus, right transverse temporal gyrus, and left precuneus, whereas the dominant negative correlations are exhibited by the right cuneus, left superior parietal lobule, right supramarginal gyrus, left occipital fusiform gyrus, and right occipital fusiform gyrus. For the AD group, the key positive correlations are portrayed by the right pallidum, third ventricle, left fusiform gyrus, right fornix, and right thalamus proper, while the key negative correlations are linked to the left subcallosal area and left frontal operculum.

**Figure 4:**
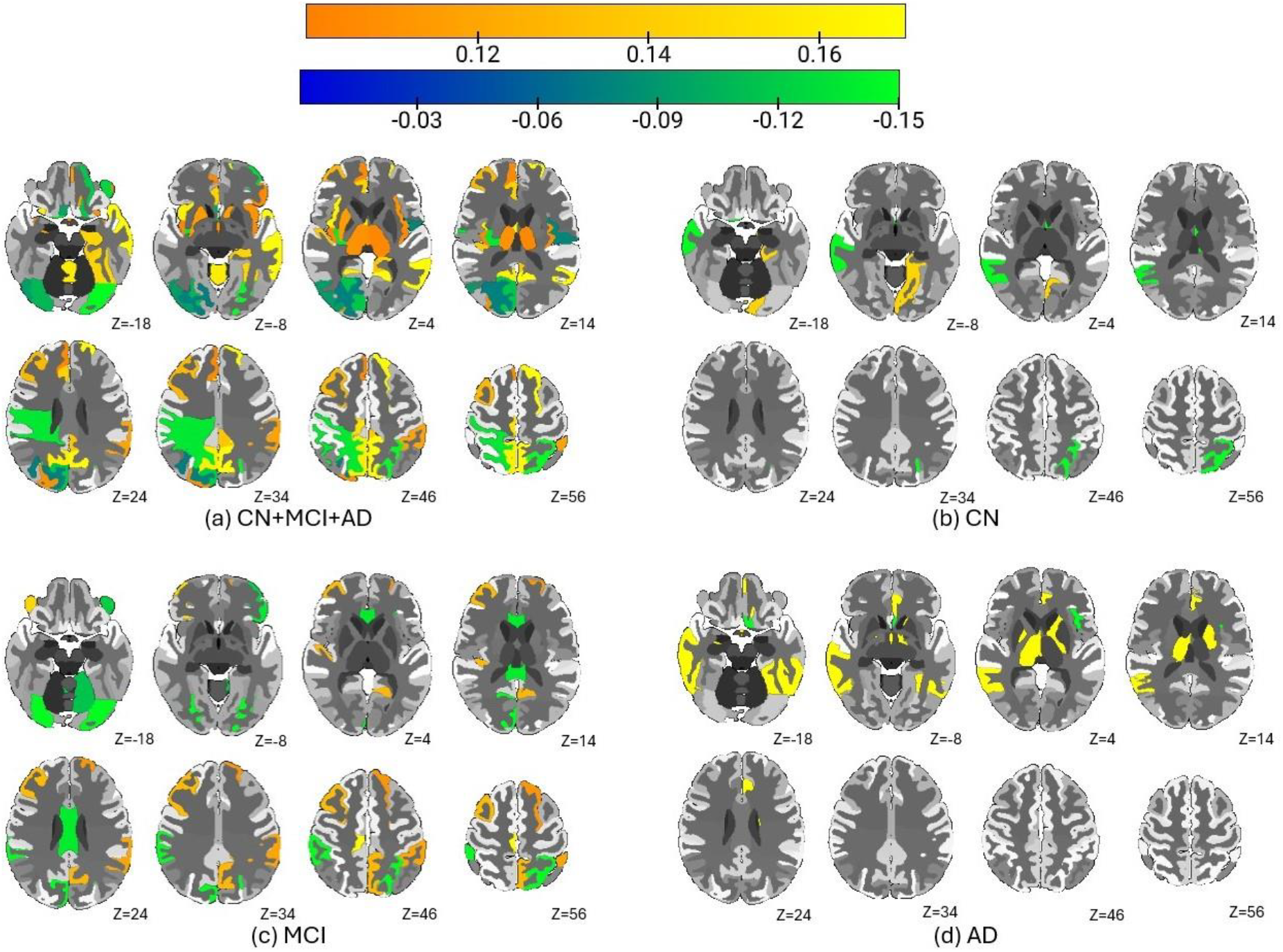
Spearman’s correlations between the individualized AgeNet-SHAP regional features and the CDR-SB scores across **(a)** Combined, **(b)** CN, and **(c)** MCI, and **(d)** AD groups. The color bar highlights the correlation values. The results are averaged over all participants. z shows the brain slice position. The gray areas show the background and are not linked with the analysis.

## 4. Discussion

This study systematically evaluated the performance of multiple ML/DL models for brain age prediction in normal aging and MCI/AD population and found that the DL model (AgeNet) outperformed the conventional ML models in both semi-simulated and experimental datasets. It further integrated the outperforming AgeNet model with SHAP feature importance strategy, and this integrated AgeNet-SHAP method captured the multivariate associations between brain age and neuroanatomical changes in CN, MCI, and AD individuals. Group differences assessed using multivariate regional AgeNet-SHAP features between the CN and MCI groups, as well as between the CN and AD groups revealed that the disease progression (i.e., from CN to MCI to AD dementia stages) leads to increased and widely distributed multivariate brain atrophy patterns. Additionally, the AgeNet-SHAP features further correlated with the participants clinical severity scores, suggesting the potential clinical relevance of these novel multivariate features.

We noticed an overlap among the most significant brain regions identified by AgeNet-SHAP across the CN, MCI, and AD groups. However, the order and magnitude of the significance varied among the groups. For instance, the third ventricle showed greater significance in AD compared to MCI and CN, while the significance of the left planum polare reduced from MCI to AD. This might suggest that left planum polare is more susceptible to AD in the early stage of the disease course. Likewise, the distinct regional hubs in MCI/AD compared to CN suggest that those regions are highly significant for AD progression. This disease progression induces brain alterations in such a way that there is a noticeable difference between brain regions and brain age, and our model effectively captures these changes. Understanding such structural brain changes at different stages of the disease course is critical for developing more targeted treatments for patients with different levels of clinical severity.

Moreover, the AgeNet-SHAP method identified several significant brain regions associated with brain age estimation, and many of our findings align with previous studies. For instance, the third ventricle was recognized as a key region for brain age in the MCI and AD groups by [33], and our method similarly highlighted it in these groups. Additionally, brainstem atrophy has been shown to correlate with aging in AD participants by [34], and our approach also linked this region to aging in AD. The left planum polare has been associated with a high brain age gap in AD [35], and AgeNet-SHAP identified it as a dominant region for brain age estimation in both MCI and AD groups. The consistency between our findings and the existing literature supports the validity of our method, confirming the relevance of the novel multivariate regions identified for brain age estimation.

Furthermore, the significant brain regions captured in the clinical severity analysis are also consistent with findings from existing literature. Like, prominent atrophies are known to happen in temporal lobe due to AD [36, 37] and our analysis also identified the left middle temporal gyrus and left temporal pole regions to have significant positive correlations with CDR-SB scores. Similarly, brain atrophy with normal aging was observed in the occipital and parietal lobes [38], and our method also identified the superior parietal lobule and occipital fusiform gyrus regions to have notable negative correlations with the CDR-SB. These findings demonstrate that AgeNet-SHAP-derived multivariate regional features are meaningful and can effectively capture clinical severity patterns associated with both the AD and normal aging processes. It is important to note that the associations between AgeNet-SHAP features and CDR-SB exhibited relatively small effect sizes, a characteristic commonly observed in brain-behaviour mappings [39].

In conclusion, this study emphasized the MRI-based modelling of the multivariate associations between brain age and brain regions by integrating the optimal AI predictive model with the feature importance technique, SHAP. It demonstrated that multiple brain regions collaboratively contribute to aging processes, and the proposed novel multivariate approach captured those significant dominant regional hubs involved in brain aging processes and clinical severity prediction. Overall, our explainable AI-based modelling approaches effectively captured multivariate regional brain age and clinical severity patterns in MCI/AD relative to CN, thereby providing a deeper understanding of disease progression mechanisms. This work can be extended in future by applying it to different neuroimaging modalities, like PET, EEG, DTI etc, which could uncover multiple brain aging aspects, aiding in more comprehensive understanding of brain aging mechanisms in MCI/AD.

## Data Availability

All data produced in the present study are available upon reasonable request to the authors

https://adni.loni.usc.edu/

